# Covid-19 rapid test by combining a random forest based web system and blood tests

**DOI:** 10.1101/2020.06.12.20129866

**Authors:** Valter Augusto de Freitas Barbosa, Juliana Carneiro Gomes, Maíra Araújo de Santana, Clarisse Lins de Lima, Raquel Bezerra Calado, Claúdio Roberto Bertoldo Júnior, Jeniffer Emidio de Almeida Albuquerque, Rodrigo Gomes de Souza, Ricardo Juarez Escorel de Araújo, Ricardo Emmanuel de Souza, Wellington Pinheiro dos Santos

**Affiliations:** Department of Mechanical Engineering, Federal University of Pernambuco, Recife, Brazil; Politechnique School of the University of Pernambuco, Recife, Brazil; Centre for Informatics, Federal University of Pernambuco, Recife, Brazil; Secretaria Municipal de Saúde de Paudalho, Paudalho, Brazil; Department of Biomedical Engineering, Federal University of Pernambuco, Recife, Brazil; Redera Technologies, Brazil

**Author notes:** Wellington Pinheiro dos Santos Email address (Wellington Pinheiro dos Santos).

**Keywords:** Covid-19, Blood tests, Software-based rapid test, Computer-aided diagnosis, Machine Learning for diagnosis, Covid-19 rapid test

## Abstract

**Background:** The disease caused by the new type of coronavirus, the Covid-19, has posed major public health challenges for many countries. With its rapid spread, since the beginning of the outbreak in December 2019, the disease transmitted by SARS-Cov2 has already caused over 400 thousand deaths to date. The diagnosis of the disease has an important role in combating Covid-19.

**Objective:** In this work, we propose a web system, Heg.IA, which seeks to optimize the diagnosis of Covid-19 through the use of artificial intelligence.

**Method:** The main ideia is that healthcare professionals can insert 41 hematological parameters from common blood tests and arterial gasometry into the system. Then, Heg.IA will provide a diagnostic report. It will indicate if the patient is infected with SARS-Cov2 virus, and also predict the type of hospitalization (regular ward, semi-ICU, or ICU).

**Results:** We developed a web system called Heg.IA to support decision-making regarding to diagnosis of Covid-19 and to the indication of hospitalization on regular ward, semi-ICU or ICU. This application is based on decision trees in a Random Forest architecture with 90 trees. The system showed to be highly efficient, with great results for both Covid-19 diagnosis and to recommend hospitalization. For the first scenario we found average results of accuracy of 92.891% ± 0.851, kappa index of 0.858 ± 0.017, sensitivity of 0.936 ± 0.011, precision of 0.923 ± 0.011, specificity of 0.921 ± 0.012 and area under ROC of 0.984 ± 0.003. As for the indication of hospitalization, we achieved excellent performance of accuracies above 99% and more than 0.99 for the other metrics in all situations.

**Conclusion:** By using a computationally simple method, based on the classical decision trees, we were able to achieve high diagnosis performance. Heg.IA system may be a way to overcome the testing unavailability in the context of Covid-19. We also expect the system will provide wide access to Covid-19 effective diagnosis and thereby reach and help saving lives.

## 1. Introduction

A highly connected world, in which physical distances among countries have been virtually reduced by modern ways of transportation, like airplanes, is also a more fragile world, from an epidemiological point of view. The ways by which the international commerce flows are the same ways used by vectors of infectious diseases. SARS and MERS are diseases disseminated by the coronaviruses SARS-Cov and MERS-Cov, respectively. They were responsible for critical outbreaks in 2002 and 2012, in this order, transmitting diseases whose main symptoms were respiratory. In December 2019, started perhaps the most critical outbreak of the recent hundred years: the rapid widespread of the coronaviruses disease 2019 (Covid-19), transmitted by SARS-Cov2, causing one of the biggest health crisis in decades. At the time of writing, SARS-Cov2 infection has resulted in over 7.5 million cases and more than 400 thousand deaths.

The accurate diagnosis has an important role against the disease. The best accepted test to diagnosis Covid-19 is the Reverse Transcription Polymerase Chain Reaction (RT-PCR) with DNA sequencing and identification (Döhla et al., 2020). However RT-PCR procedures takes several hours (Döhla et al., 2020). The late diagnosis might imply in late patient care and making it difficult the patient recovery. Besides that non-isolated infected people could spread the virus.

The most common rapid tests are basing in identify serological evidences of the virus presence as antibodies or antigens. Thus those such tests are nonspecific for not detecting the virus presence directly. So rapid tests might response positively to Covid-19 from samples containing other coronavirus (that cause the common cold) or flu virus (Li et al., 2020; WHO, 2020). That is why the tests performance depends of factors like the time from onset of illness, the concentration of virus in specimen WHO (2020). For example, tests based on IgM/IgG antibodies realized in the first week of illness have 18.8% of sensitivity and 77.8% of specificity (Liu et al., 2020b). However, in the second week the IgG/IgM tests perform with 100% of sensitivity and 50% of specificity (Liu et al., 2020b). So when the viral charge is high IgG/IgM tests reach high sensitivities and specificities. But in that case the disease is in advanced levels (Guo et al., 2020; Hoffman et al., 2020). Because of that the World Health Organization does not recommend the use of this kind of tests for clinical decision-making WHO (2020).

Many studies have been evidencing that Covid-19 affect the blood (Fan et al., 2020; Tan et al., 2020; Gao et al., 2020; Liu et al., 2020a; Gunčar et al., 2018; Zheng et al., 2020). The coronaviruses, like SARS-Cov and SARS-Cov2, have as functional receptor the zinc metallopeptidase angiotensin-convertingenzyme 2 (ACE2), an enzyme presents in cell membranes of arteries, heart, lungs and other organs. ACE2 is implicated in heart function, hypertension and diabetes (Turner et al., 2004). Middle East respiratory syndrome-related coronavirus (MERS-CoV) and severe acute respiratory syndrome coronavirus (SARS-Cov) can cause acute myocarditis and heart failure (Zheng et al., 2020). According to Zheng et al. (2020) some of the coronaviruses impacts in cardiovascular system are the increase in blood pressure, and increase in troponin I (hs-cTnI) levels (Zheng et al., 2020). Besides that patients with Covid-19 can developed lymphopenia (low level of lymphocytes in the blood) (Fan et al., 2020; Tan et al., 2020; Liu et al., 2020a), leukopenia (few white blood cells). They can also have decrease in their hemoglobin levels, Absolute Lymphocyte Count (ALC) and Absolute Monocyte Count (AMC) (Fan et al., 2020). Patients that developed severe forms of the disease have significantly higher levels of hematological characteristics as Interleukin-6, D-dimer than patients that developed moderate form of the Covid-19 (Gao et al., 2020). Therefore, considering that Covid-19 is a disease that affects blood parameters, hematological tests can be used to diagnosis the disease.

Machine learning, a branch of the Artificial Intelligence in Computer Science, is the area that studies techniques specialized in pattern detection. Machine learning techniques have been used to many pattern detection tasks as image classification (Lerner et al., 1994; Phung et al., 2005; Barbosa et al., 2020b; Gomes et al., 2020b), image reconstruction (Gomes et al., 2020a), biosignals analyzing (Müller et al., 2008; Karlik, 2014; Jambukia et al., 2015; Andrade et al., 2020), hematological parameters analyzing (Tanner et al., 2008; Luo et al., 2016; Gunčar et al., 2018) and etc.

In this work, we propose a web diagnosis support system of the Covid-19 based in machine learning techniques. This system uses blood tests to diagnose Covid-19. Our machine learning method were training using the database provided by Hospital Israelita Albert Einstein located in São Paulo, Brazil. The database is formed by information from 5644 patients among them 559 patients were diagnosis with Covid-19 by RT-PCR with DNA sequencing and identification and additional laboratory tests during a visit to the hospital (Kaggle, 2020). For each patient the database have more than one hundred laboratory tests like blood counts and urine test. From this database we set a new one that contains only 41 blood tests recommended by the Brazilian Ministry of Health when dealing with Covid-19 patients. Our goal is to provide a web tool that perform accurate Covid-19 diagnosis with friendly interface and low computational cost.

## 2. Related works

Several studies have shown evidence of the relationship between Covid-19 and the blood. Moreover, they emphasize the importance of blood tests for the diagnosis process of this disease. There are also studies that point to the relevance of using hematological analysis as an indicative of the severity degree of Covid-19. Fan et al. (2020) analyzed hematological parameters of 69 patients with Covid-19. The study was conducted with subjects from the National Center for Infectious Diseases (NCID) in Singapore. 65 of these patients underwent complete blood count (CBC) on the day of admission. 13.4% of patients needed intensive care unit (ICU) care, especially the elderly. During the first exams, 19 patients had leukopenia (few white blood cells) and 24 had lymphopenia (low level of lymphocytes in the blood), with 5 cases classified as severe (Absolute Lymphocyte Count (ALC) < 0.5 × 10^9^*/L*). The study also pointed out that patients who needed to be admitted to the ICU had lower ALC and a higher rate of Lactate Dehydrogenase (LDH). These data indicated that monitoring these parameters can help to identify patients who need assistance in the ICU. The authors found that the patients who were in the ICU had a significant decrease in their hemoglobin levels, ALC and Absolute Monocyte Count (AMC) levels, when compared to the non-ICU group. ICU patients also tend to neuthophilia. The platelet count did not prove to be a factor for discrimination between the type of hospitalization.

The work from Tan et al. (2020) also assessed the complete blood count of patients. They used data from both cured patients and patients who died from Covid-19. The main purpose of the study was to obtain key indicators of disease progression, in order to support future clinical management decisions. In the case of patients who died, blood tests were continuously monitored throughout the treatment process. Similar to the previous study, the authors observed lymphopenia in this group. Based on this, the study then outlined a model (Time-LYM% model, TLM) for classifying disease severity and predicting prognosis. Thus, the blood lymphocyte percentage (LYM%) was divided into two cases, considering the first 10-12 days of symptoms: LYM% > 20% are classified as moderate cases and with a high chance of recovery. LYM% < 20% are classified as severe cases. In a second exam, 17-18 days after the first symptoms, patients with LYM% > 20% are recovering, patients with 5 <LYM% < 20% are in danger, and LYM% < 5% are in critical condition. In order to validate the model, the authors evaluated 90 patients with Covid-19. The consistency between Guideline and TLM-based disease classification was verified using kappa statistic (Kappa = 0.48). These results indicate that probably LYM% should be used together with other parameters for a better evaluation of Covid-19.

In Gao et al. (2020), the authors assessed hematological characteristics of 43 patients at Fuyang Second People’s Hospital. The patients had diagnosis confirmed by the Covid-19 ground truth test, the fluorescent reverse transcription-polymerase chain reaction (RT-PCR). They were divided into two groups: the moderate group with 28 patients, and the severe group with 15 patients. The groups have no significant difference in age and sex. The blood tests observed were: Routine blood tests (white blood cell [WBC] count, lymphocyte count [LYM], mononuclear count [MONO], neutrophils count [NEU]) were performed on the blood samples. Blood biochemistry parameters (aspartate aminotransferase [AST], alanine aminotransferase [ALT], glucose [GLU], urea, creatinine [Cr], cystatin [Cys-c], uric acid [UA], and C-reactive protein [CRP]); Coagulation functions (the D-dimer [d-D], thrombin time [TT], prothrombin time [PT], fibrinogen [FIB], activated partial thromboplastin time [APTT]); rocalcitonin (PCT); and Interleukin-6 (IL-6). Using statistical tests, the study noted that the levels of GLU, CRP, IL-6, TT, FIB, and d-D were significantly higher in the severe group than in the mild group. Performing this analysis with ROC curves, the authors pointed out that the best indicators for predicting severity were IL-6 and d-D combined, with AUC of 0.840. The combination also achieved specificity of 96.4% and sensitivity of 93.3%, using tandem and parallel testing, respectively. These results indicate that patients with severe conditions would have abnormal coagulation.

Liu et al. (2020a) reported that lymphopenia and inflammatory cytokine storm are abnormalities commonly found in other infections caused by coronavirus, such as SARS-Cov and MERS-Cov. With that in mind, they studied 40 patients diagnosed with Covid-19 confirmed by throat-swab specimens analyzed with RT-PCR. The patients were treated at Wuhan Union Hospital in January, 2020. The information provided was: epidemiological, demographic, clinical manifestations and laboratory tests. Similar to the previous study, patients were divided into two groups: mild patients, with symptoms such as epidemiological history, fever or respiratory symptoms, and abnormalities in imaging tests; the second group with severe patients, patients should additionally have symptoms such as shortness of breath, oxygen saturation < 93%, respiratory > 30 times/min, or PaO_2_*/*FiO_2_ < 300 mmHg. 27 patients were classified in the first group, while 13 were classified in the second. The study reported that levels of fibrinogen, D-dimer, total bilirubin, aspartate transaminase, alanine transaminase, lactate dehydrogenase, creatine kinase, C-reactive protein (CRP), ferritin and serum amyloid A protein were significantly higher in the severe group. Futhermore, most severe patients presented lymphopenia, that can be related to the significantly decreased absolute counts of T cells, especially CD8+ T cells, while white blood cells and neutrophils counts were higher.

These studies have pointed out that hematological parameters can be indicators of the risk factors and degree of severity of Covid-19. The identification of these parameters can be essential to optimize clinical care for each group of patients. In this sense, the development of intelligent systems based on blood tests is useful. Faced with the pandemic scenario, in which most hospitals are full, decision support systems can facilitate clinical management. Thus, it can increase the assertiveness in the treatment for each case and, consequently, the number of lives saved.

Gunčar et al. (2018) proposed a system based on machine learning for analyzing blood tests and predicting hematological diseases. Their database was acquired between the years 2005 and 2015 at the University Medical Center of Ljubljana. In this case, 43 diseases and 181 parameters or features were selected to generate a first model (SBA-HEM181). In addition to it, a second model with 61 parameters was also developed (SBA-HEM061). The selection criteria was based on the frequency of use. Regarding the missing values (about 75%), the authors filled in with median values for each attribute. As classification methods, the authors tested classic approaches, such as Support Vector Machines, Naive Bayes and Random Forest. The simulations were repeated 10 times using 10-fold cross validation. Finally, the models SBA-HEM181 and SBA-HEM061 reached an accuracy of 57% considering all the diseases chosen. By restricting the prediction to five classes, the systems achieved an accuracy of 88% and 86%, respectively. These results were achieved when using Random Forest for classification. This study also pointed to the possibility of effectively detecting diseases through blood tests using classic intelligent classifiers.

In our previous work (Barbosa et al., 2020a) we proposed an intelligent system to aid Covid-19 diagnosis using blood exams. After testing several machine learning methods, we achieved high classification performance using Bayes network as classifier. Our system was built using a public database from the Hospital Israelita Albert Einstein, in Brazil (Kaggle, 2020) and showed average accuracy of 95.159% ± 0.693, with kappa index of 0.903 ± 0.014, sensitivity of 0.968 ± 0.007, precision of 0.938 ± 0.010 and specificity of 0.936 ± 0.011. In this study we were able to minimize costs by selecting only 24 blood tests from the set of 107 available exams. However, although we managed to achieve good results, the set of selected tests did not considered all the exams indicated by subsequent recommendations from the Brazilian Ministry of Health when dealing with Covid-19 patients (Brazilian Ministry of Health, 2020).

Similarly, the study by Soares et al. (2020) uses a method based on artificial intelligence to identify Covid-19 through blood tests. As in our previous work (Barbosa et al., 2020a), they used the database from the Hospital Israelita Albert Einstein. However, since the database has many missing data, they chose to include only the subjects that had most of the data. This procedure reduced the dataset from 5,644 samples to 599 samples. By using Support Vector Machines as a classifier and SMOTEBoost technique to perform oversampling, they achieved average specificity of 85.98%, negative predictive value (NPV) of 94.92%, average sensitivity of 70.25% and positive predictive value (PPV) of 44.96%.

These intelligent systems based on blood tests may play an important role in the process of diagnosing Covid-19, since many studies are confirming evidences of this disease on blood. Moreover, as previously mentioned, most of these exams are simple, fast and widely available. Based on this initial classification using blood tests, positive cases can be referred to further highly sensitive testing such as RT-PCR with virus DNA identification, CT scan and Radiography.

In this sense, many other studies are also being conducted in order to optimize Covid-19 diagnosis using these other testing methods. Gomes et al. (2020c) proposed a new technique for representing DNA sequences to optimize the molecular diagnosis of Covid-19. Their method divides the DNA sequences into smaller sequences with overlap in a pseudo-convolutional approach, and represented by co-occurrence matrices. The DNA sequences are obtained by the RT-PCR method, eliminating sequence alignment. Through this approach, it is possible to identify virus sequences from a large database by using AI tools to improve both specificity and sensitivity. The group conducted experiments with 347,363 virus DNA sequences from 24 virus families and SARS-Cov-2. They used three different scenarios to diagnose SARS-Cov-2. In the first scenario the authors used all 24 families and SARS-Cov-2. They achieved 0.822222 ± 0.05613 for sensitivity and 0.99974 ± 0.00001 for specificity when using Random Forests with 100 trees and 30% overlap. For the second scenario they aimed to compare SARS-Cov-2 with similar-symptoms virus families. In this condition, MLP classifier with 30% overlap performed better, showing sensitivity of 0.97059 ± 0.03387 and specificity of 0.99187 ± 0.00046. Finally, in the third scenario, they tested the real test scenario, in which SARS-Cov-2 is compared to Coronaviridae and healthy human DNA sequences. For this last condition they found 0.98824 ± 001198 for sensitivity and 0.99860 ± 0.00020 for specificity with MLP and 50% overlap.

Another study from our group (Gomes et al., 2020b) shows benefits of using artificial intelligence to perform automatic detection of Covid-19 in X-ray images. In this work, we proposed an intelligent system able to differentiate Covid-19 from viral and bacterial pneumonia in X-ray images, with low computational cost. There are many advantages of using X-rays images in Covid-19 diagnosis process: it is a widely available technique, with low cost, and fast delivery time. In this work we tested Haralick and Zernike moments for feature extraction. After representing these images, we conducted experiments with several classical classifiers. Among the tested configurations, we found that Support Vector Machines overcame the other methods, reaching an average accuracy of 89.78%, average recall and sensitivity of 0.8979, and average precision and specificity of 0.8985 and 0.9963, respectively.

## 3. Materials and methods

### 3.1. Proposed method

In this work we present an intelligent system in web format called Heg.IA. Heg.IA is a system that seeks to optimize the diagnosis of Covid-19 through the use of artificial intelligence. The basic idea is that healthcare professionals can log in to the system and register patients in care. Then, it is possible to insert the results of blood tests that are commonly requested for patients with characteristic symptoms of Covid-19. After completing the filling, the professional can view and print the report. The report will indicate from the blood tests if the patient is infected with the SARS-Cov2 virus. In addition, it will also indicate whether the patient should be admitted to the regular ward, Semi-intensive care unit (Semi-ICU) or Intensive care unit (ICU). These results will be accompanied by the values of accuracy, sensitivity, specificity and kappa index, helping the physician in making the final decision. The diagram in the Figure 1 shows a summary of this solution.

**Figure 1:**
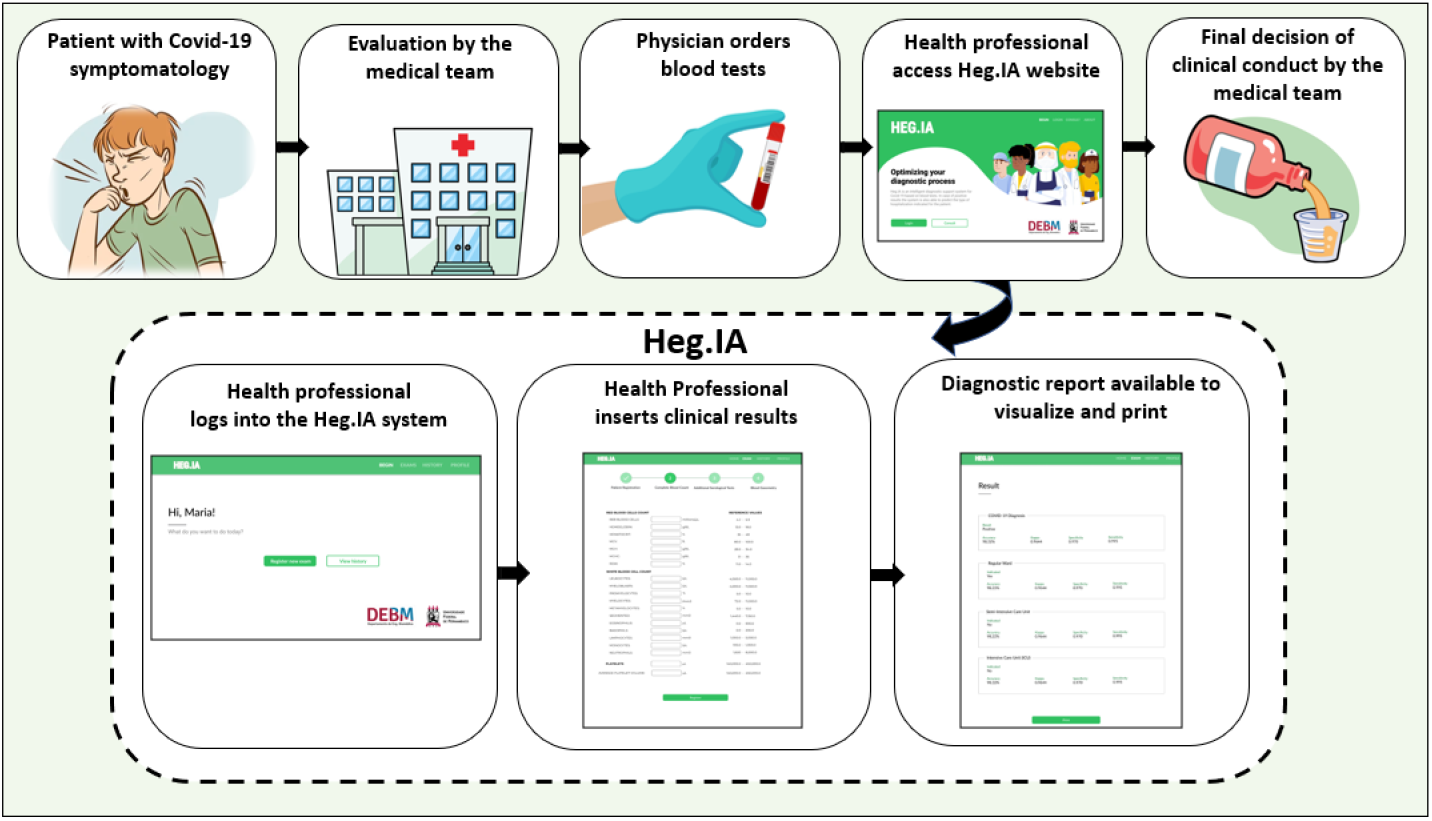
General proposed method: The main idea is for a patient with symptoms characteristic of Covid-19 to go to a health center. He will be evaluated by a medical team, who must order blood tests. After obtaining the results, a health professional will be able to access the HegIA website. On the website, he must log in. Then he can enter the patient’s blood test results. After finishing, the system will generate a report with a positive or negative diagnosis for Covid-19, in addition to the hospitalization prediction. This report can be printed and used for the medical team to define the final clinical conduct.

It is important to note that health professionals actively participated in the process of developing the system’s front-end. Thus, the interface of the developed system is easy to use and does not require long periods of user training. Furthermore, the developed back-end uses conventional classifiers. These choices made it possible to achieve a low computational cost, and to make the result available almost immediately.

In addition to the functionality of registering patients, the website also allows access by the other physicians who have not participated in the blood test analysis process. This access will be through a patient’s private locator. For this type of user, only the report’s visualization and printing functions are available. These system use cases are described in the diagram in the Figure 2.

**Figure 2:**
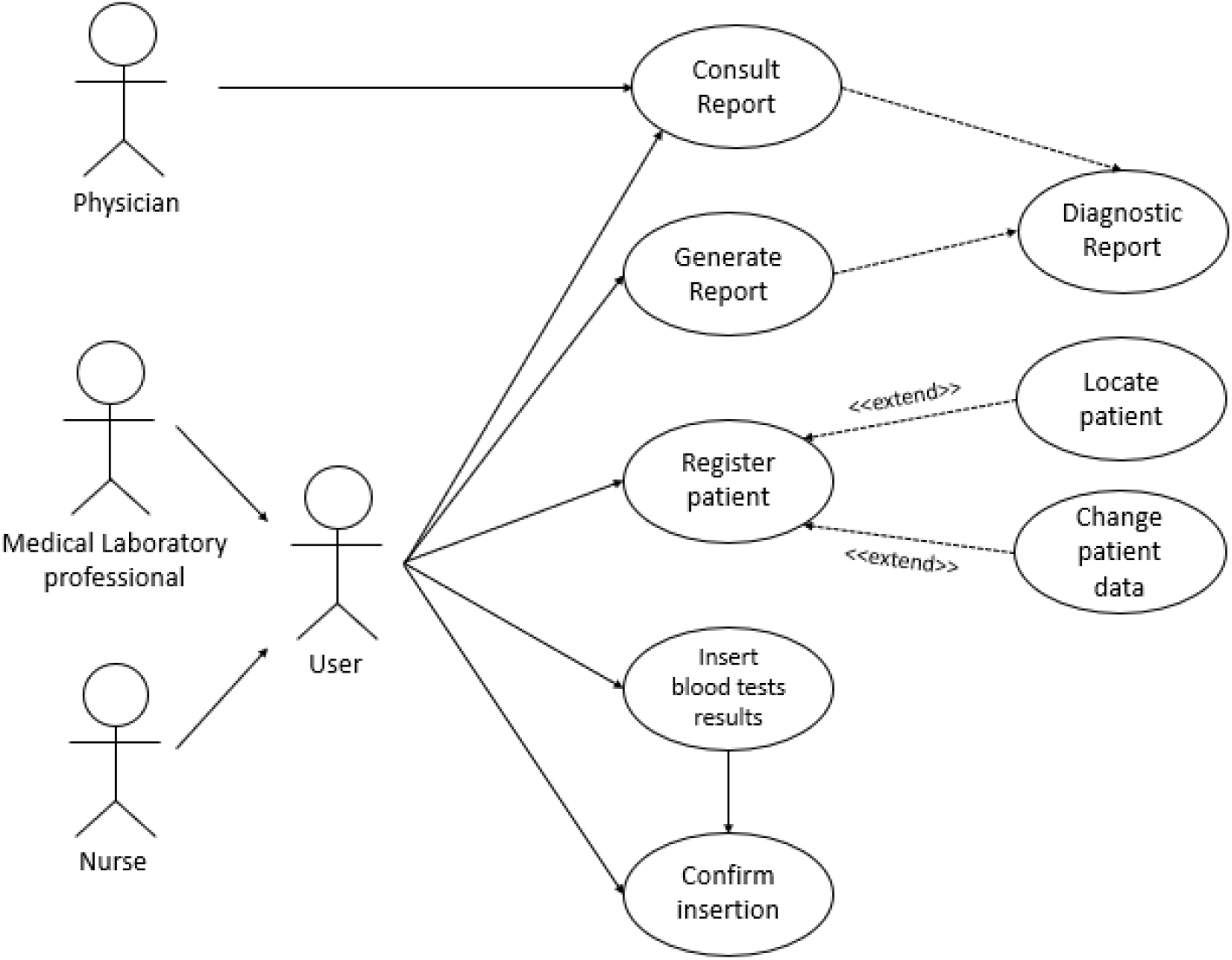
Use case Diagram: The main users of the system are nurses and medical laboratory professionals. They will be able to register patients, insert patients’ blood test results and check if the data was entered correctly. They can also generate diagnostic reports and consult them at any time. Physicians will be able to view the report, accessing the system through a patient’s personal locator.

### 3.2. Database

In this work, we used a public database with information on hospitalized patients at Hospital Israelita Albert Einstein, located in São Paulo (Brazil) (Kaggle, 2020). The database consists of data from 5644 patients with symptoms similar to Covid-19. All patients’ personal information was omitted from the database, respecting privacy, with the exception of the age. Patients underwent multiple clinical examinations, including blood tests, arterial and venous blood gas analysis, urine tests and rapid tests for various types of viruses. The exams total 107 clinical parameters. In addition, all patients were tested for Covid-19 by analyzing swabs by RT-PCR with DNA sequencing, the current gold standard for the disease. Among the patients, 559 tested positive for Covid-19. The database also presented information regarding the hospitalization of each patient, indicating whether they were treated in the regular ward, semi-intensive care unit or in the ICU.

### 3.3. Database Pre-processing

In the database, not all 5644 patients underwent all types of clinical examinations. Therefore, the base has several missing values. In addition, several exams or attributes are categorical, that is, they have nominal results (or classes). As the objective of this work is to use this data as input parameters for machine learning methods, it was necessary to handle the missing data and transform the categories into numerical classes. The transformations made are described in the following Table 1. The left column indicates the attributes that have been modified, while the right column indicates the numerical values assigned to each category. For the case of the result of pathological tests for SARS-Cov2, for instance, it was assigned the value 0 for not detected cases, 1 when abnormalities are detected, and 2 for missing values.

**Table 1:**
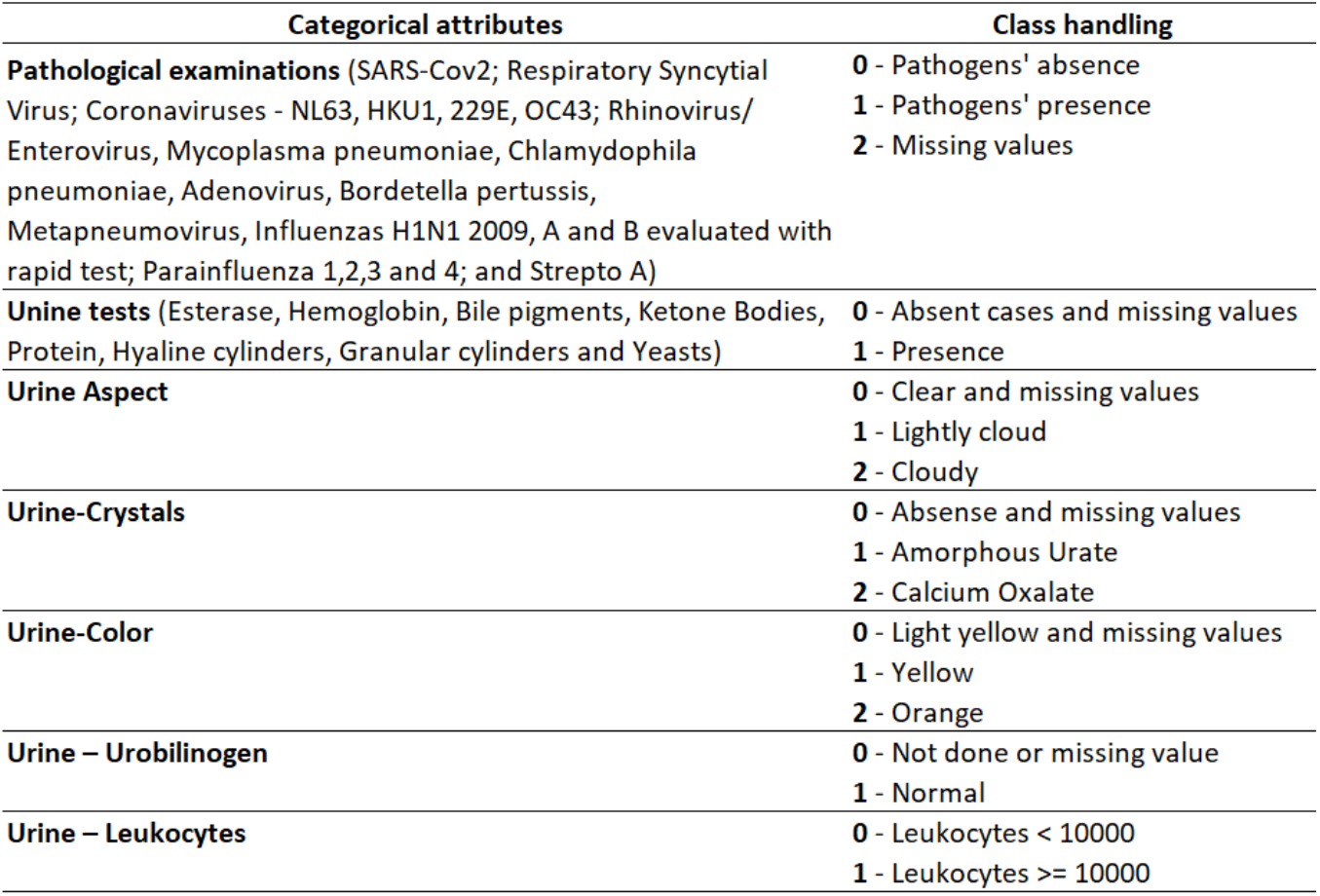
Database pre-processing: The attributes with categorical classes were reorganized, receiving numerical values corresponding to each of the classes. The attributes related to pathological exams, such as SARS-Cov2, types of influenza and parainfluenza, received the values of 0, 1 and 2 for the absence of pathogens, presence of pathogens, and missing values, respectively. The attributes “Unine tests”, “Urine Crystals”, “Unire color”, “urine urobilinogen” and “urine leukocytes” also have been modified, according to the labels indicated in right column.

Finally, it was also necessary to treat the missing data from the columns with numerical classes. In these cases, a value of zero has been assigned. As the base is normalized with a mean of zero and standard deviation of 1, this transformation means that the missing data were filled in with the average value of each of the exams. This assumption is valid, since 90.1% of the patients in the database have a negative result for Covid-19, and therefore, their parameters may be considered normal.

### 3.4. Exams Selection

Among the 107 exams available in the initial database, 41 exams were selected. The exams chosen correspond to those recommended by the Ministry of Health of Brazil as an initial clinical approach and part of the Covid-19 diagnostic process (Brazilian Ministry of Health, 2020). Thus, considering that health centers must already perform these tests, there is no financial loss or time spent on additional tests. On the contrary, the diagnostic process can be optimized with the system proposed here.

The list of 41 hematological parameters is shown in the Figure 3. The Complete Blood Count (CBC) with differential comprises 20 of these parameters, while arterial blood gas analysis includes 9 parameters. The remaining 12 exams are those of total, indirect and direct Bilirubin; Serum Glucose; Lipase dosage; Urea; D-Dimer; Lactic Dehydrogenase; C-Reactive Protein (CRP); Creatinine; and Partial thromboplastin time (PTT) and Prothrombin time Activity from coagulogram.

**Figure 3:**
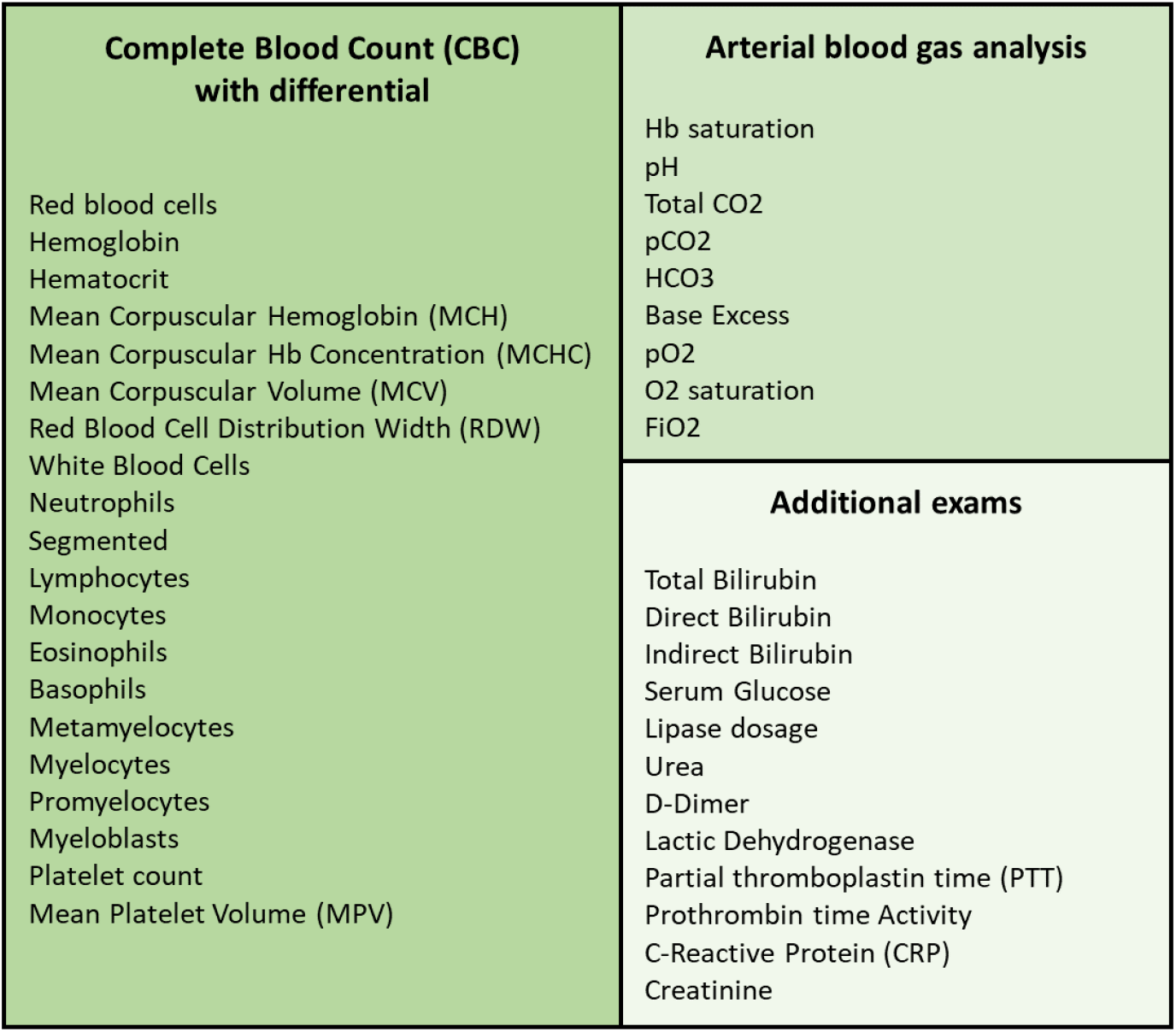
List of exams

### 3.5. Classification

#### 3.5.1. Multilayer Perceptron

Multilayer perceptron (MLP) consists of a generalization of the Perceptron proposed by Franklin Rosenblatt in 1958. Perceptron is the model is the simplest form of a neural network, being able to deal with linearly separable problems. Multilayer Perceptron networks, on the other hand, have several interconnected neurons (or nodes), arranged in layers: the input layer, the hidden layers and the output layer. The input layer only has the network input vector, which is passed on to the next layer. Then, each node in the next layer modifies these input values through non-linear activation functions, generating output signals. In addition, the network nodes are connected by weights, which scales these output signals. Finally, the superposition of several non-linear functions allows the mapping of the input vector to the output vector. As MLPs can have one or multiple hidden layers, this process can be repeated several times, depending on the selected architecture (Gardner & Dorling, 1998; Lerner et al., 1994; Phung et al., 2005; Barbosa et al., 2020b).

Thus, through the proper selection of activation functions and synaptic weights, an MLP is able to approximate the inputs at the desired outputs. This search and adjustment of parameters is called the training process. MLPs learn in a supervised manner. During this process, errors between the actual and desired outputs are calculated. These errors are used to adjust the network (Gardner & Dorling, 1998).

In order to adjust these weights, the backpropagation algorithm is the most computationally straightforward and common algorithm. It occurs in two phases: the forward and backward propagation. In the first step, the initial network weights are set to small random values. Then, this first input vector is propagated through the network to obtain an output. This actual output is compared with the desired one, and the error is calculated. In the second phase, the backward propagation, the error signal is propagated back through the network and the connection weights are updated, aiming to minimise the overall error. These steps can be repeated until the overall error is satisfactory (Haykin, 2001).

MLPs and other artificial neural networks architectures are commonly used in support diagnosis applications (Naraei et al., 2016), e.g. liver disease dianogis (Abdar et al., 2018), heart diasese diagnosis (Hasan et al., 2017), breast cancer diagnosis over breast thermography (de Vasconcelos et al., 2018; Pereira et al., 2020b; Santana et al., 2020; Pereira et al., 2020a,c; Santana et al., 2018; Rodrigues et al., 2019) and mammography images (de Lima et al., 2016; Lima et al., 2015; de Lima et al., 2014; Silva et al., 2020; Cordeiro et al., 2017, 2016; de Lima et al., 2014; Cruz et al., 2018), for recognition of intracranial epileptic seizures (Raghu & Sriraam, 2017), and multiple sclerosis diagnosis support (Commowick et al., 2018).

#### 3.5.2. Support Vector Machine

Support Vector machines (SVM) were created by Vladimir Vapnik and Alexey Chervonenkis (Boser et al., 1992; Cortes & Vapnik, 1995) in 1963. Their main purpose is to build a linear decision surface, called a hyperplane. The idea is that this hyperplane should be able to separate classes in the best possible way. The optimal hyperplane is found when the margin of separation between it and a given nearest point is maximum (Haykin, 2001).

SVM is known for its good generalization performance. Therefore, it is used in several healthcare applications, such as breast cancer diagnosis using thermography and mammography (de Vasconcelos et al., 2018; Pereira et al., 2020b; Santana et al., 2020; de Lima et al., 2016; Lima et al., 2015; de Lima et al., 2014; Silva et al., 2020; Cordeiro et al., 2017, 2016; de Lima et al., 2014; Cruz et al., 2018), diabetes mellitus diagnosis (Barakat et al., 2010), heart valve diseases (Çomak et al., 2007) and pulmonay infections detection (Yao et al., 2011), and also diagnosis of pulmonary cancer (Sun et al., 2013). However, its performance varies depending on the problems complexity. The type of the machine varies with the type of kernel used to build the optimal hyperplane. Table 2 shows the kernel functions used in this study: the polynomial and RBF kernels. For the first case, it was tested exponents of value 1 (linear kernel), 2, and 3.

**Table 2:** Kernel functions of SVM

| Type of SVM | Kernel function |
| --- | --- |
| Polynomial | $K(\mathbf{x}, \mathbf{y}) = (\mathbf{x} \bullet \mathbf{y} + 1)^E$ |
| Radial Basis Function (RBF) | $K(\mathbf{x}, \mathbf{y}) = \exp(-\gamma (\mathbf{x} - \mathbf{y}) \bullet (\mathbf{x} - \mathbf{y}))$ |

#### 3.5.3. Decision Trees

Decision trees are sequential models, which combine several simple tests. They can be understood as a series of questions with “yes” and “no” answers. These tests can be the comparison of a value with a threshold or a categorical attribute compared to a set of possibilities, for instance. Thus, when analyzing the data with these tests, the decision trees will guide to a certain class in classification problems, or to a continuous value, in cases of regression problems. In this way, a decision tree is built with certain questions, called nodes. Essentially, there area four types of nodes: root, parent, child, and leaf. Starting at the root node, an instance is classified. Then, the outcome for this instance is determined ad the process continues through the tree. In addition, one node may connect to another, establishing a parent-child relationship, in which a parent node generate a child node. Finally, the terminal nodes of the tree are the leaf nodes, and they represent the final decision, that is, the predicted class or value. There are several types of decision trees, depending on the tree structure. The most popular ones are Random Tree and Random Forest. Both of them were tested in this study by using multiple parameters (Kotsiantis, 2013; Podgorelec et al., 2002).

Random Tree uses a tree built by a stochastic process. This method considers only a few randomly selected features in each node of the tree Geurts et al. (2006). In contrast, Random Forest is a model made up of many decision trees. In this case, a set of trees is built and their votes are combined to classify an instance, by means of the majority vote. Each decision tree uses a subset of attributes randomly selected from the original set of attributes Breiman (2001).

#### 3.5.4. Bayesian Network and Naive Bayes

Bayesian classifiers are based on Bayes’ Decision Theory. Among the most popular Bayesian classifiers are Naive Bayes and Bayes Net. Bayesian networks describe the probability distribution over a set of variables. They represent, in a simple way, the causal relationships of the variables of a system using Graph Theory, where the variables are the nodes and the arcs identify the relationships between the variables. In the learning process, it is necessary to calculate the probability distributions and to identify the network structure. Learning the network structure can be considered an optimization problem, where the quality measure of a network structure needs to be maximized Cheng & Greiner (1999); Bouckaert (2008).

On the other hand, the Naive Bayes classifier is a simple model that considers that the domain variables are conditionally independent, that is, one characteristic is not related to the other. Its learning is done in an inductive way, presenting a set of training data and calculating the conditional probability of each attribute, given a class. Naive Bayes needs to estimate few parameters (Cheng & Greiner, 2001; Bouckaert, 2008).

### 3.6. Parameters settings of the classifiers

All experiments were performed using the Weka software. The experiments were made by using the following techniques: SVM with polynomial kernel of degree (*E*) 1, 2, and 3 and RBF kernel with *γ* of 0.01; MLP with 50 and 100 neurons in hidden layer; Random Forest with 10, 20, 30, …, 100 trees; Random Tree; Naive Bayes; Bayesian Network. As a evaluate method we choose to perform a 10-fold cross validation and each configuration was executed 25 times.

### 3.7. Metrics

We chose seven metrics to evaluate the performance of diagnostic tests: accuracy, precision, sensitivity, specificity, recall, precision and the area under ROC. Accuracy is the probability that the test will provide correct results, that is, be positive in sick patients and negative in healthy patients. In other words, it is the probability of the true positives and true negatives among all the results. The recall and sensitivity metrics can be calculated mathematically in the same way. They are the rate of true positives, and indicate the classifier ability to detect correctly people with Covid-19. However, they are commonly used in different contexts. In machine learning context, the term Recall is common. However, in the medical world, the use of the sensitivity metric is more frequent. Precision, on the other hand, is the fraction of the positive predictions that are actually positive. Specificity is the capacity of classifying healthy patients as negatives. It is the rate of true negatives. The Kappa index is a very good measure that can handle very well both multi-class and imbalanced class problems, as the one proposed here.

Finally, the area under the ROC curve is a measure of a classifier’s discriminating ability. That is, given two classes a sick individual and a non-sick individual -, chosen at random, the area below the ROC curve that indicates a probability of the latter being correctly classified. If the classifier can not discriminate between these two separately, an area under a curve is equal to 0.5. When this value is the next 1, it indicates that the classifier is able to discriminate these two cases (Hand, 2009).

These metrics allow to discriminate between the target condition and health, in addition to quantifying the diagnostic exactitude (Borges, 2016). The accuracy, precision, sensitivity, specificity, recall and precision can be calculated according to the equations in Table 3.

**Table 3:**
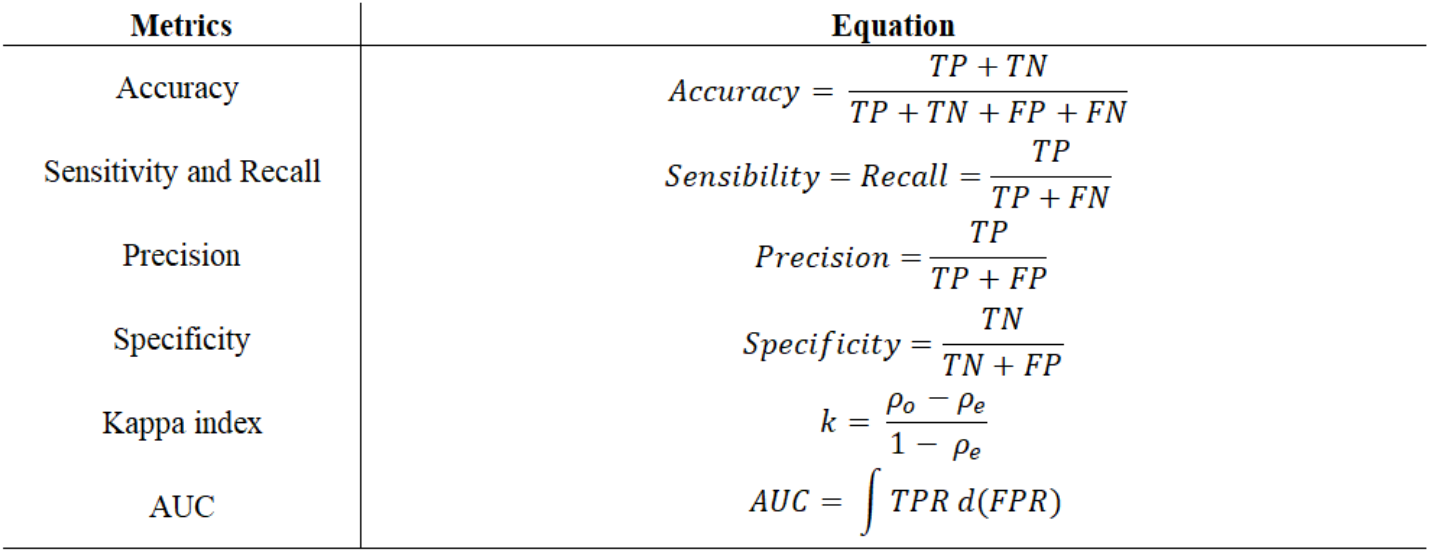
Metrics used to evaluate classifiers performance: overall accuracy, sensitivity (recall), precision, specificity, and kappa index

| Metrics | Equation |
| --- | --- |
| Accuracy | $Accuracy = \frac{TP + TN}{TP + TN + FP + FN}$ |
| Sensitivity and Recall | $Sensibility = Recall = \frac{TP}{TP + FN}$ |
| Precision | $Precision = \frac{TP}{TP + FP}$ |
| Specificity | $Specificity = \frac{TN}{TN + FP}$ |
| Kappa index | $k = \frac{\rho_o - \rho_e}{1 - \rho_e}$ |
| AUC | $AUC = \int TPR d(FPR)$ |

In Table 3, TP is the true positives, TN is the true negatives, FP is the false positives, and FN the false negatives, *ρ*_*o*_ is observed agreement, or accuracy, and *ρ*_*e*_ is the expected agreement, defined as following:

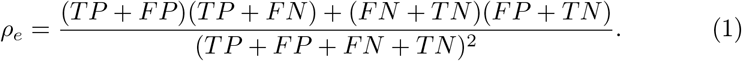

## 4. Results

This section presents the results obtained for the evaluation of different computational methods to detect SARS-Cov2 and to indicate the type of hospitalization. Furthermore, we show the main screens of our web system.

### 4.1. Model Validation

Since the proposed system aims to perform the tasks of diagnosing SARS-Cov2 and recommending the type of hospitalization, we performed the classification tests considering each scenario.

#### 4.1.1. Detection of SARS-Cov2

Figure 4 shows the results of accuracy and kappa statistic for all configurations of the studied classifiers. These results present the performance of different algorithms in diagnosing SARS-Cov2 from the 41 blood exams previously mentioned.

**Figure 4:**
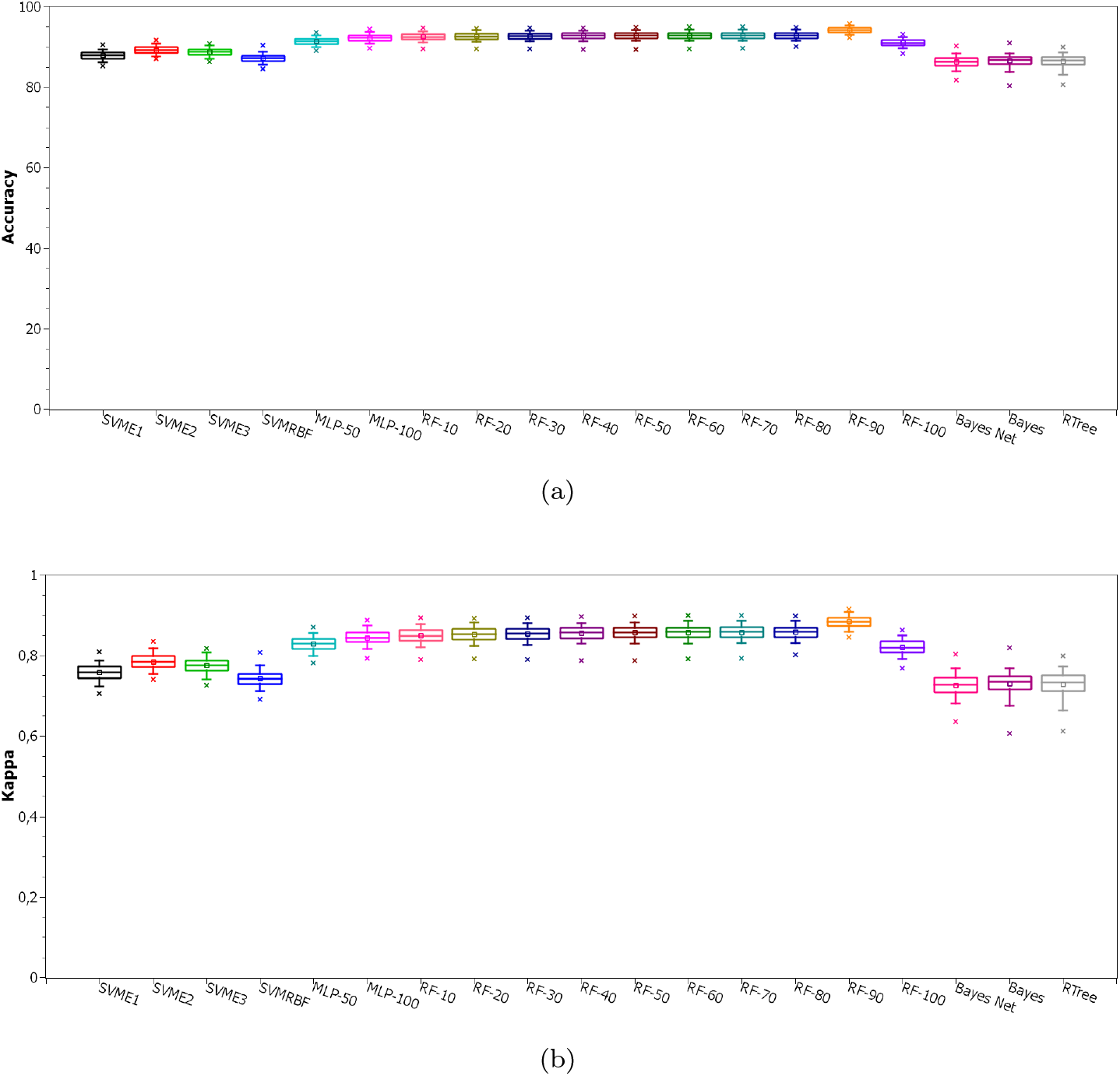
Classification performance for detection of SARS-Cov2. In (a) are the results of accuracy while (b) shows the kappa results.

In Table 4 we show the average values and standard deviation of all metrics to measure the performance of the Random Forest algorithm with 90 trees in detecting SARS-Cov2, as long as it has the higher accuracy and kappa index.

**Table 4:** Classification performance using Random Forest with 90 trees for SARS-Cov2 detection.

| Database | Accuracy (%) |  | Kappa statistic |  | Recall/Sensitivity |  | Precision |  | Specificity |  | Area under ROC |  |
| --- | --- | --- | --- | --- | --- | --- | --- | --- | --- | --- | --- | --- |
|  | Average | StdDev | Average | StdDev | Average | StdDev | Average | StdDev | Average | StdDev | Average | StdDev |
| SARS-Cov2 | 92.8914 | 0.8511 | 0.8578 | 0.0170 | 0.9365 | 0.0109 | 0.9226 | 0.0112 | 0.9213 | 0.0123 | 0.9836 | 0.0028 |

#### 4.1.2. Indication of Hospitalization

In the graphs shown in Figures 5 to 7, we present the performance of all tested classifiers to point out the recommended type of hospitalization. As mentioned before, in this scenario we have 3 possible choices to refer patients according to the severity of each case: Intensive Care Unit, Semi Intensive Care Unit or Regular Ward. Figure 5 shows the results of accuracy and kappa statistic for the Intensive Care Unit. Figures 6 and 7 present the achieved results for Semi Intensive Care Unit and Regular Ward, respectively.

**Figure 5:**
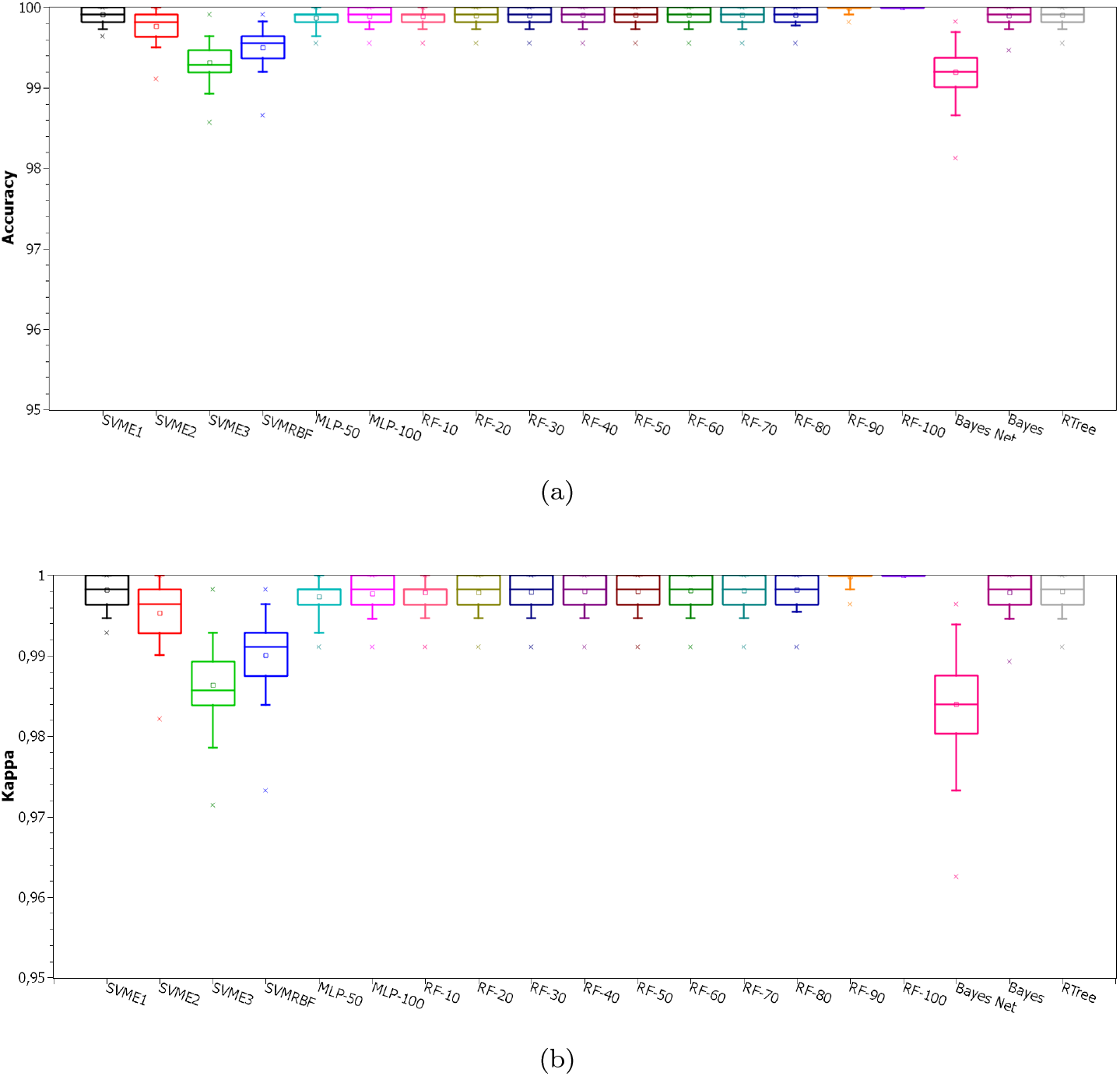
Classification performance for Intensive Care Unit indication. In (a) are the results of accuracy while (b) shows the kappa results.

**Figure 6:**
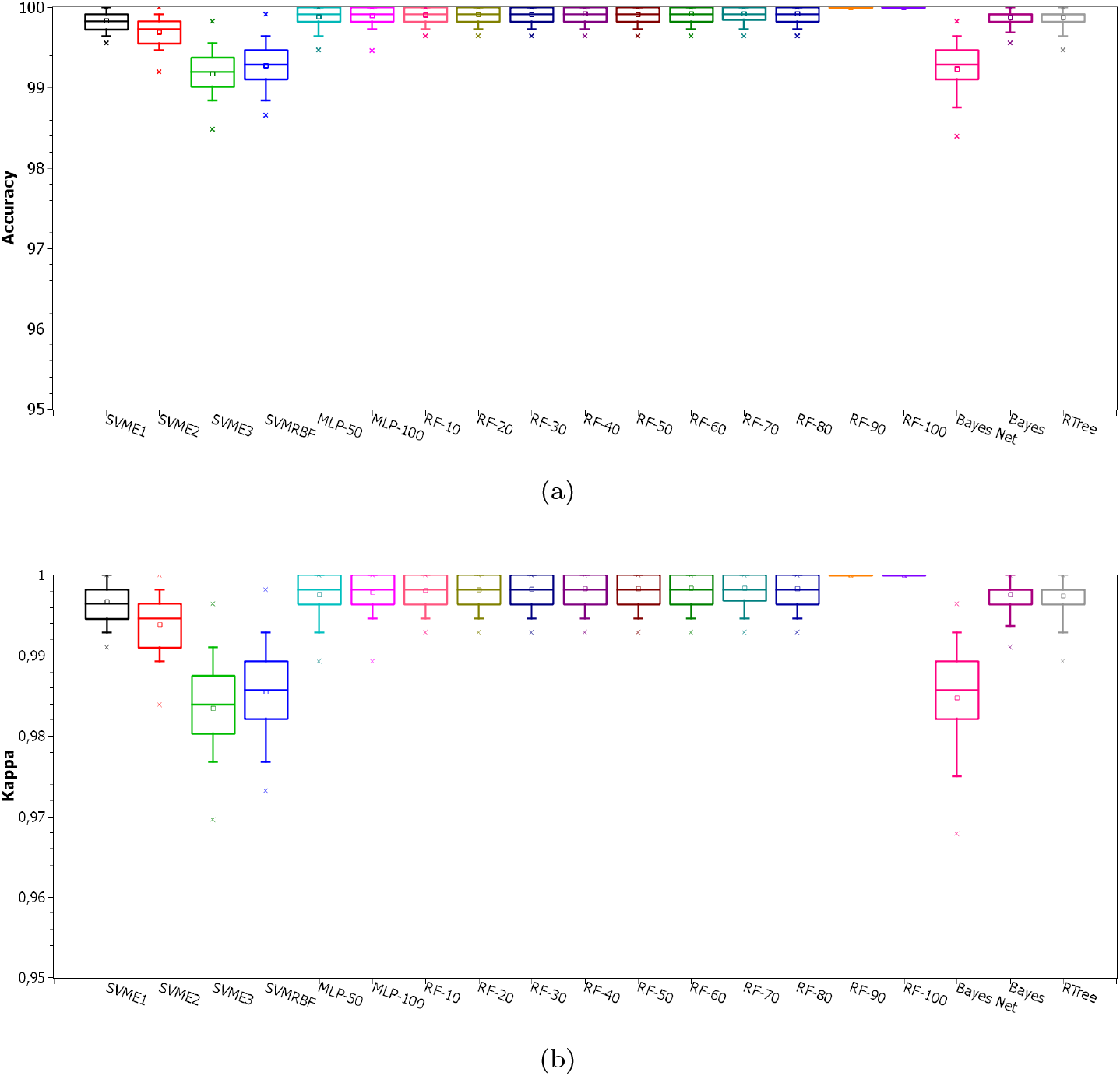
Classification performance for Semi Intensive Care Unit indication. In (a) are the results of accuracy while (b) shows the kappa results.

**Figure 7:**
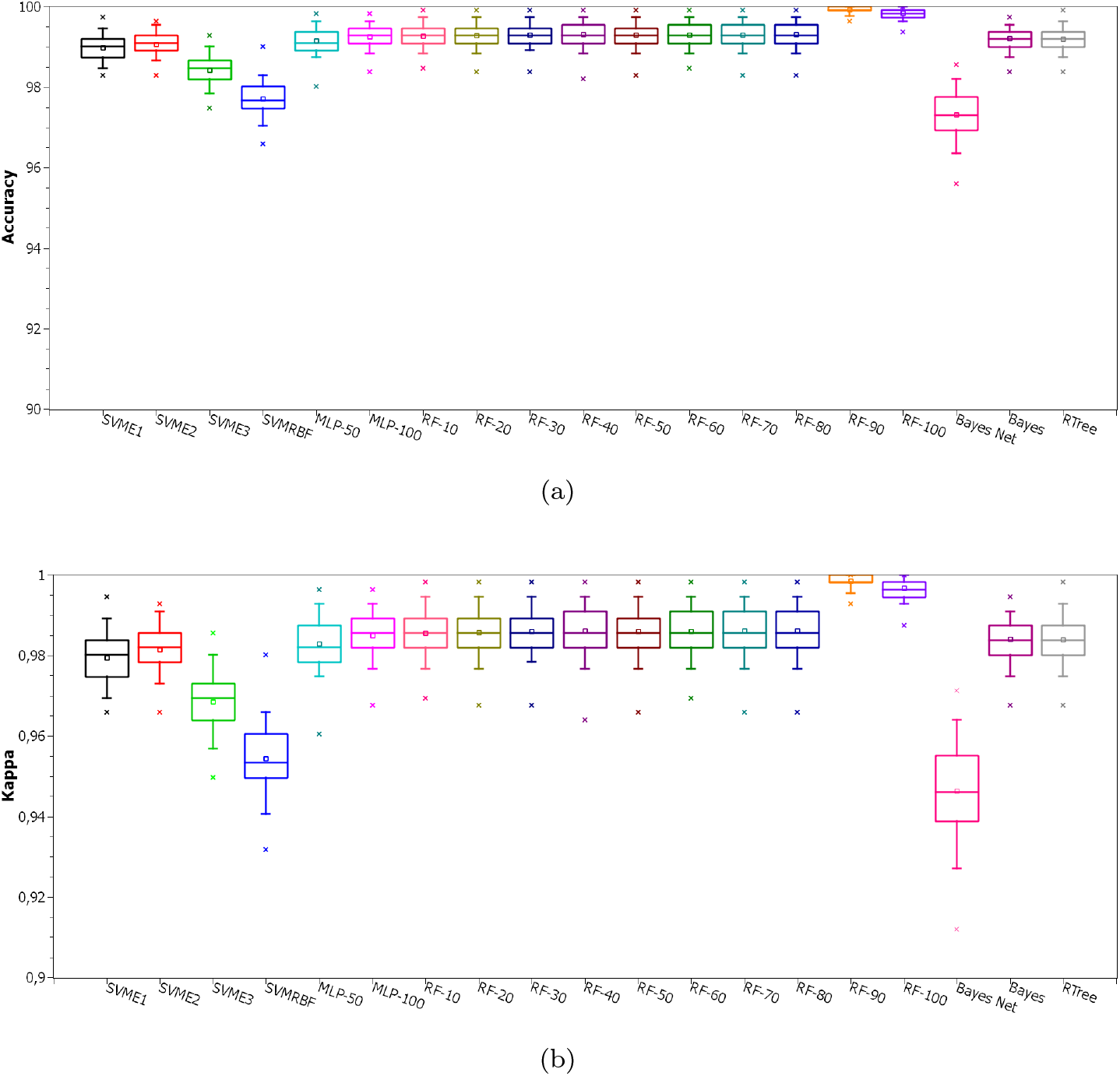
Classification performance for Regular Ward indication. In (a) are the results of accuracy while (b) shows the kappa results.

In Table 5 are the average values and standard deviation of all metrics to measure the performance of the Random Forest algorithm with 90 trees for the hospitalization prediction scenario.

**Table 5:** Classification performance using Random Forest with 90 trees for the indication of hospitalization.

| Database | Accuracy (%) |  | Kappa statistic |  | Recall/Sensitivity |  | Precision |  | Specificity |  | Area under ROC |  |
| --- | --- | --- | --- | --- | --- | --- | --- | --- | --- | --- | --- | --- |
|  | Average | StdDev | Average | StdDev | Average | StdDev | Average | StdDev | Average | StdDev | Average | StdDev |
| Intensive Care Unit | 99.9058 | 0.0826 | 0.9981 | 0.0016 | 0.9981 | 0.0016 | 0.9999 | 0.0001 | 1.0000 | 0.0001 | 0.9999 | 0.0000 |
| Semi Intensive Care Unit | 99.9196 | 0.0864 | 0.9984 | 0.0017 | 0.9989 | 0.0014 | 0.9995 | 0.0009 | 0.9995 | 0.0001 | 0.9999 | 0.0000 |
| Regular Ward | 99.3049 | 0.2748 | 0.9861 | 0.0055 | 0.9903 | 0.0046 | 0.9958 | 0.0028 | 0.9958 | 0.0028 | 0.9998 | 0.0002 |

### 4.2. HegIA Web Application

After selecting the best classifier, the Heg.IA web system was developed. It can be accessed through the link: http://150.161.141.202/welcome. Its front-end was developed using the library React.js. This library is based on pure JavaScript. It is open source and used to create user interfaces, more specifically, single page application (SPA) web platforms. As for data access and manipulation of application state, we used the Redux-Saga structure, a powerful tool that allows us to manage masterfully asynchronous queries, receiving API data, and trigger actions to the application of state safely and easily to maintain. Furthermore, our back-end was developed in Python (version 3.7.7). Only the Random Forest classifier was implemented in this final solution.

On the initial screen, as shown on Figure 8, it is possible to visualize a brief description of the intelligent system, as well as the supporters of this initiative: The Federal University of Pernambuco (UFPE) and the Department of Biomedical Engineering at UFPE. To get to know the members of the project’s development team and their respective functions, it is possible to access the ‘About’ option on the top menu of the screen. The options ‘Login’ and ‘Consult’ are also available. For the ‘Login’ option, health professionals, especially medical laboratory professionals and nurses, will be able to access their private account or register a new account, in cases of first access. In the Consult option, it is possible to view the report with the diagnosis for a specific patient, as long as the user has the patient’s personal locator.

**Figure 8:**
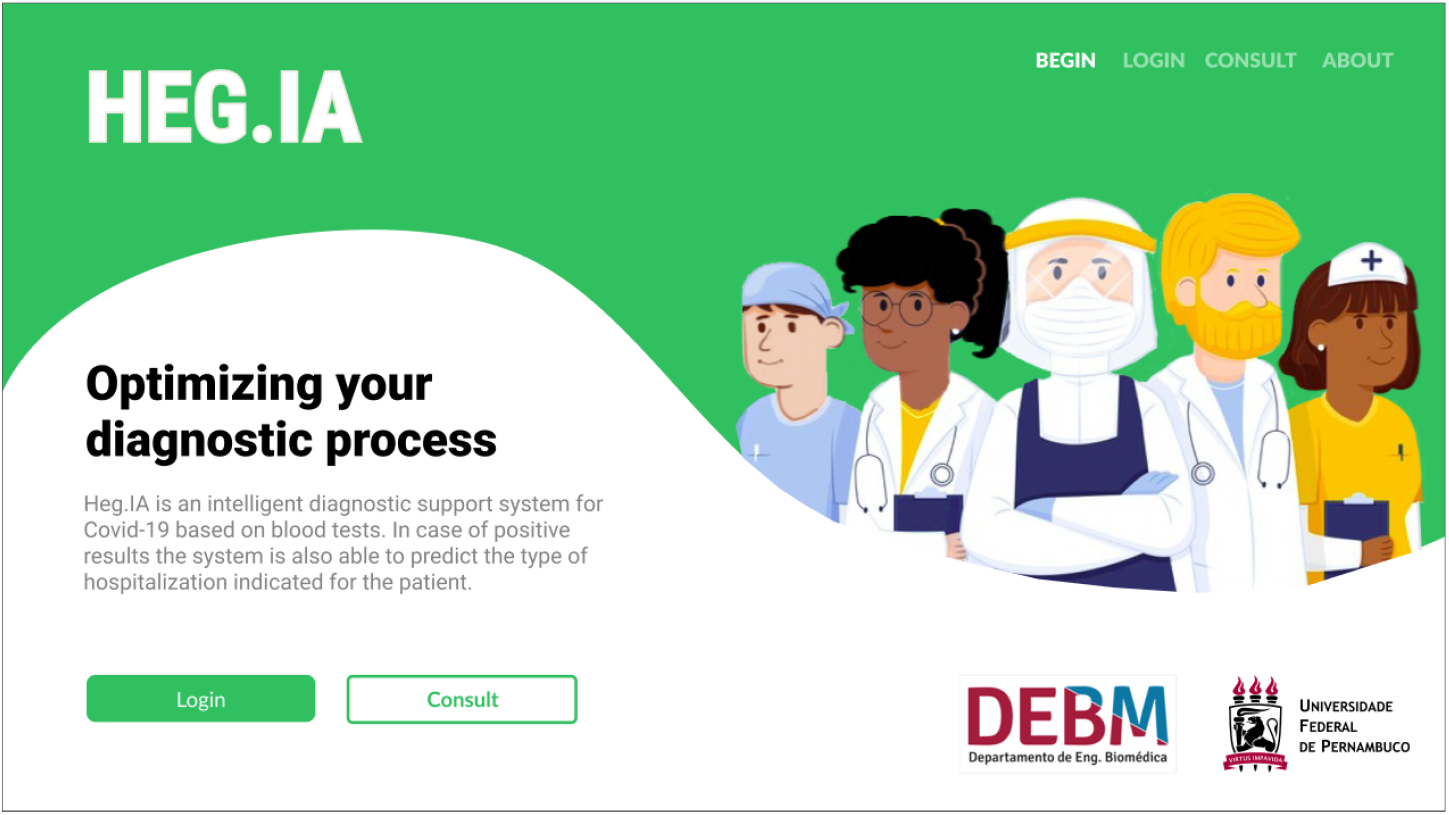
Heg.IA homepage: an intelligent web-based system for diagnosing Covid-19 through blood tests. On this initial screen, it is possible for the user (nurses and medical laboratory professionals) to login in his personal account. There are also the ‘Consult’ and ‘About’ options, available for all users, including patients and physicians. They allow the user to visualize the diagnostic report, and to get to know the involved team in this project, respectively.

After logging into the system, the user can register a patient or view the complete history of registered patients. In the case of a new registration, personal information such as full name, ID, date of birth, telephone, sex and full home address will be requested (Figure 9). In the following, the user will be directed to the screen shown in the Figure 10. In this screen, the results of the Complete blood count (CBC) with differential must be entered. The units and reference values are available next to each of the hematological parameters. After filling in the CBC, the user will be directed to the screens for the other blood tests and arterial blood gas tests, as shown in the Figures 11 and 12. Thus, the list of tests required to make the predictions will be complete. The user can then check the parameters entered in the screen ‘Let’s check it out?’, as shown in the Figure 13. If he realizes that he made a typo, he can go back to the previous steps and correct it.

**Figure 9:**
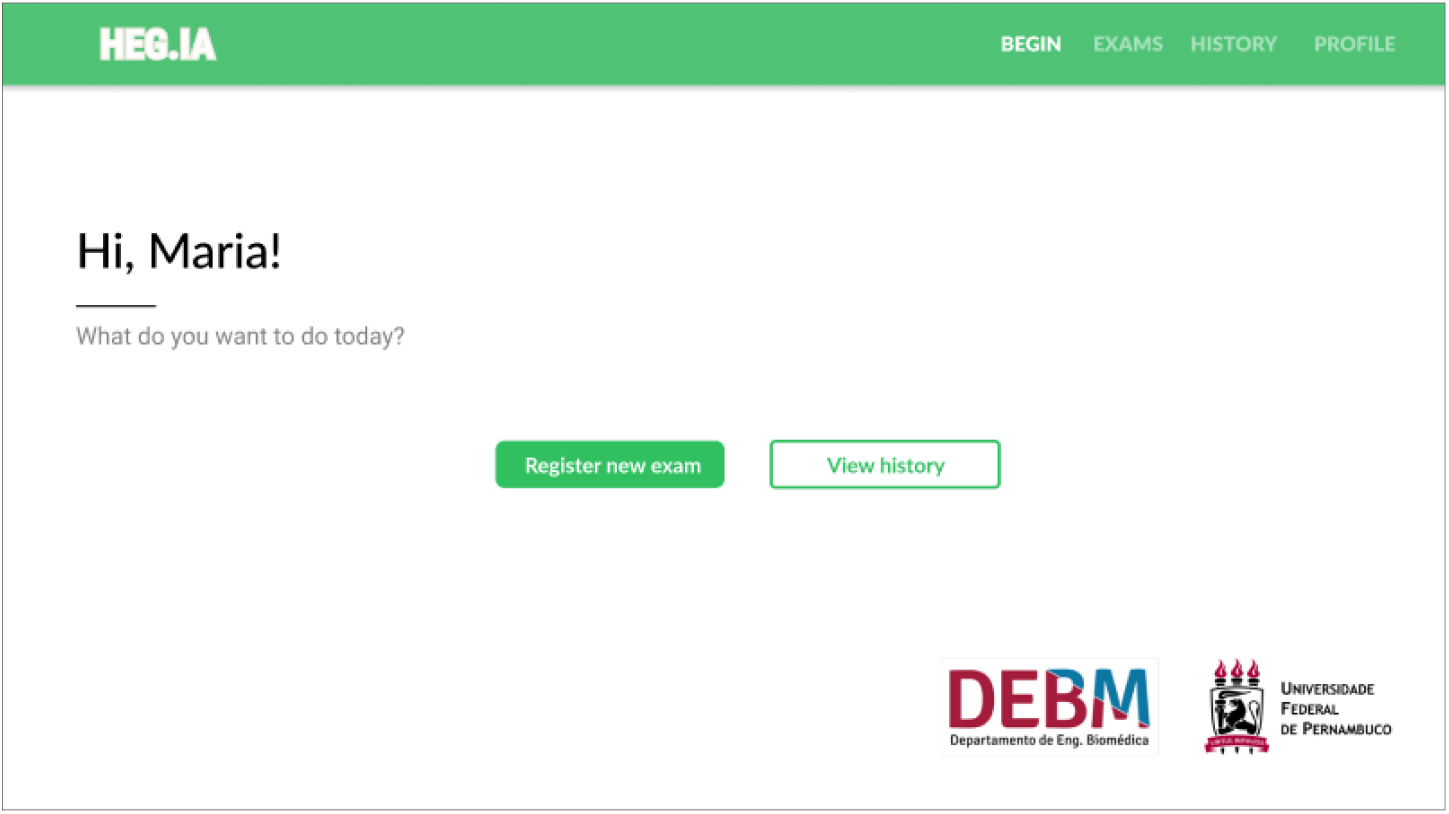
On this screen, the logged in user will be able to register new patients or access the complete history of patients already registered.

**Figure 10:**
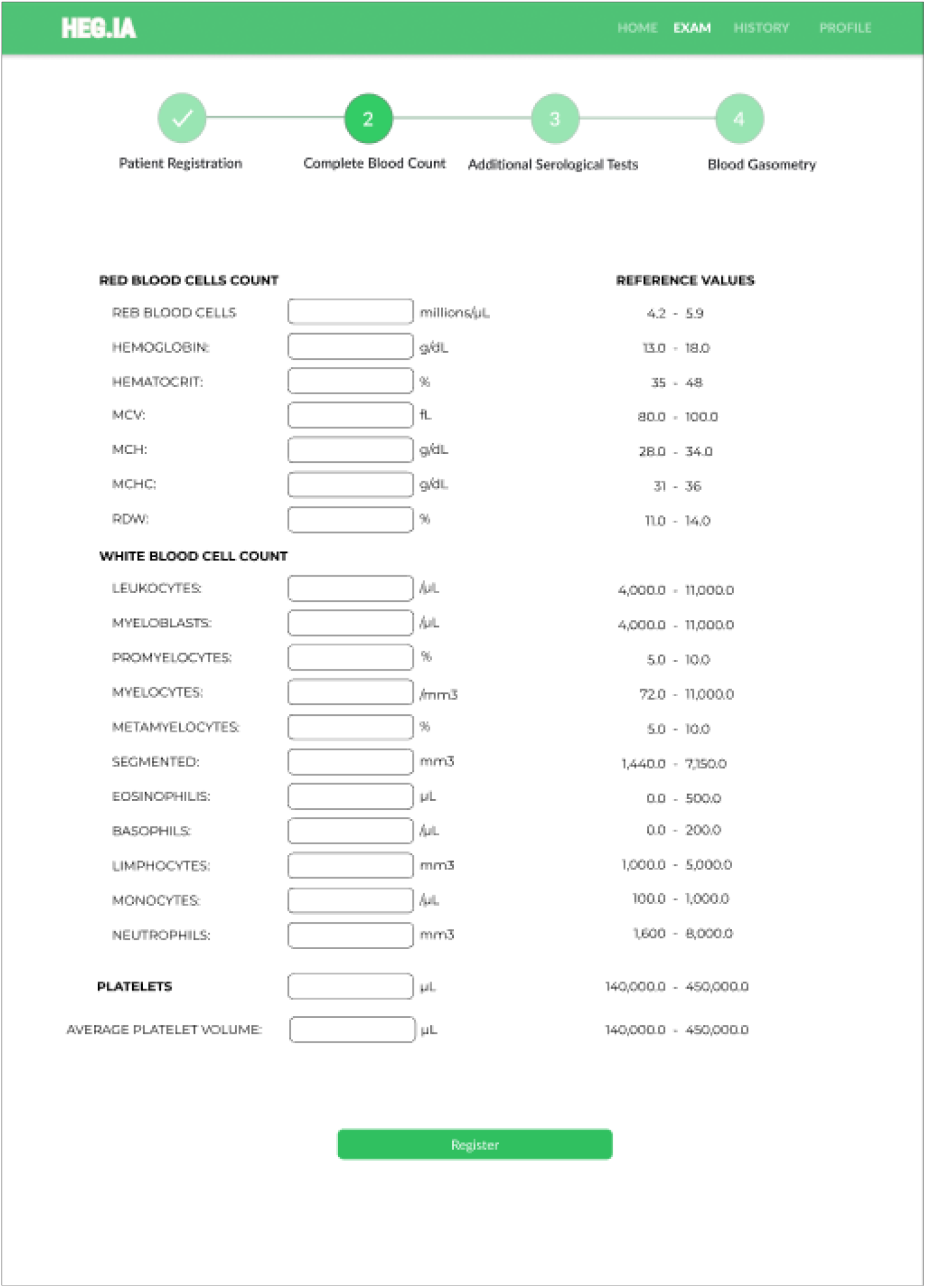
Complete Blood Count screen: After the patient’s registration, the results of the patient’s Complete Blood Count with differential can be inserted. The units and reference values can be viewed next to each hematological parameter.

**Figure 11:**
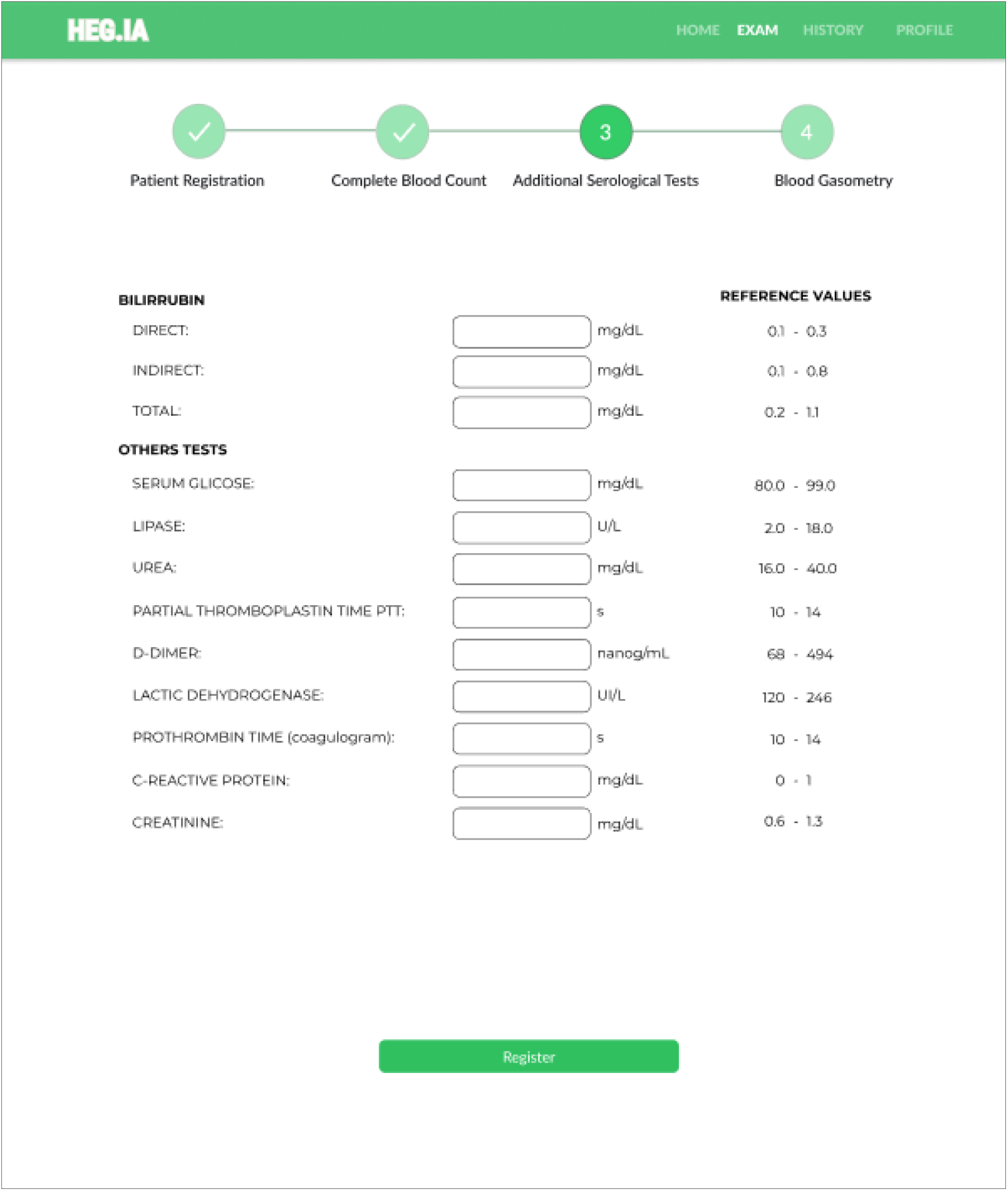
Additional Serological Tests Screen: Additional tests, such as total, direct and indirect bilirubin, can be inserted in this screen. In addition, serum glicose, dosage lipase, urea, PTT, D-dimer, lactic dehydrogenase, prothrombin time, CRP, and creatinine results can also be included here. The units and reference values can be viewed next to each hematological parameter.

**Figure 12:**
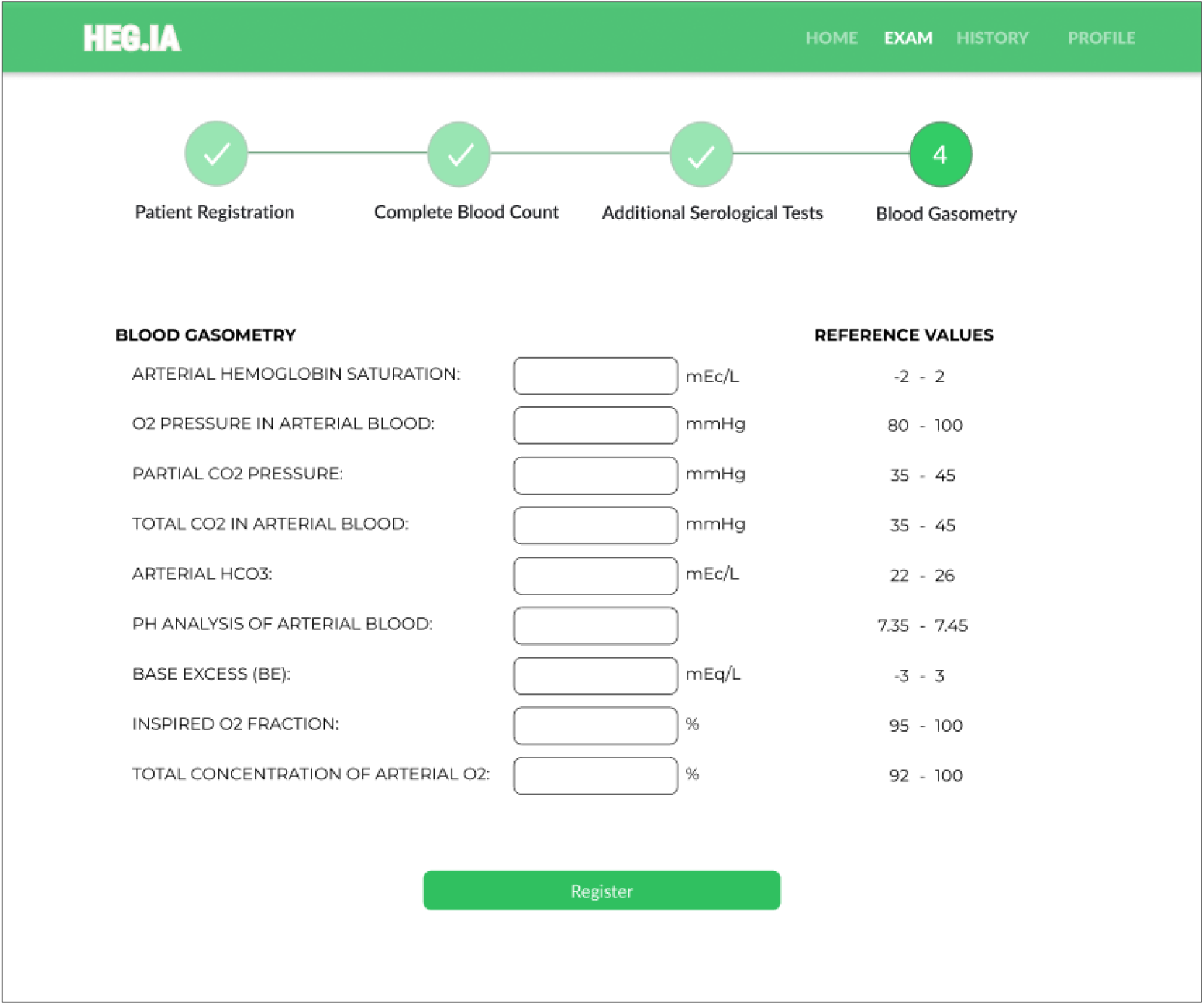
Blood Gasometry Screen: Arterial gasometry can be inserted in this screen, finalizing the list of necessary exams. The units and reference values for each parameter are available.

**Figure 13:**
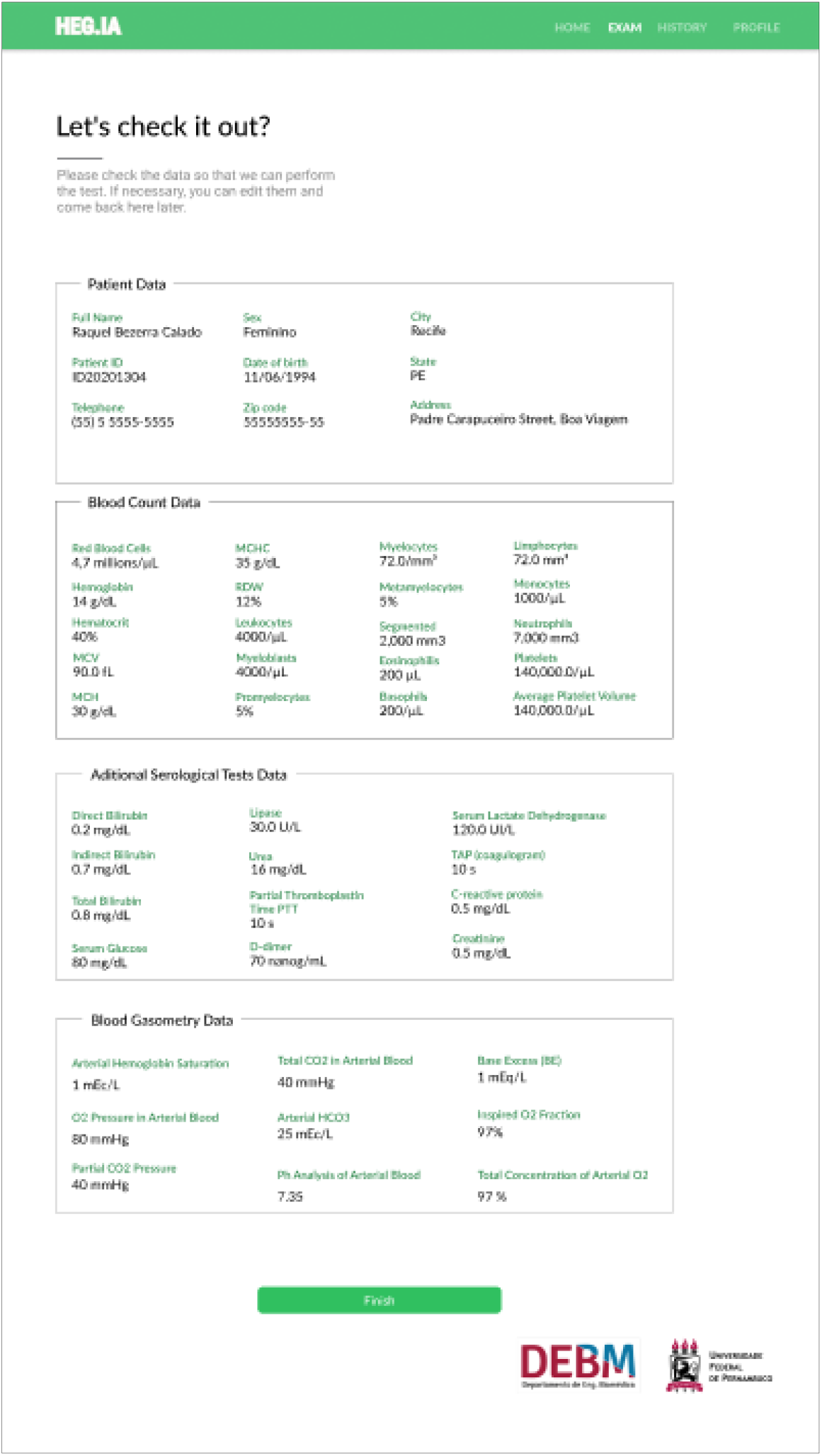
In the screen, it is possible for the user to check the patient’s personal information and the values of the hematological parameters inserted. If there is a typo, the user can return to the previous screens to correct it. If they are correct, it is possible to select the “Finish” option in order to access the report.

Finally, the report will be available immediately, similarly to that shown in the Figure 14. The diagnostic report will indicate the positive or negative diagnosis for Covid-19. Hospitalization predictions are also reported, indicating the best type of hospitalization for the patient: regular ward, semi-ICU or ICU. Information on accuracy, kappa index, sensitivity and specificity of the determination of each of these scenarios are also available, in order to assist the physician’s decision making. In addition to viewing the report, it is also possible to print it.

**Figure 14:**
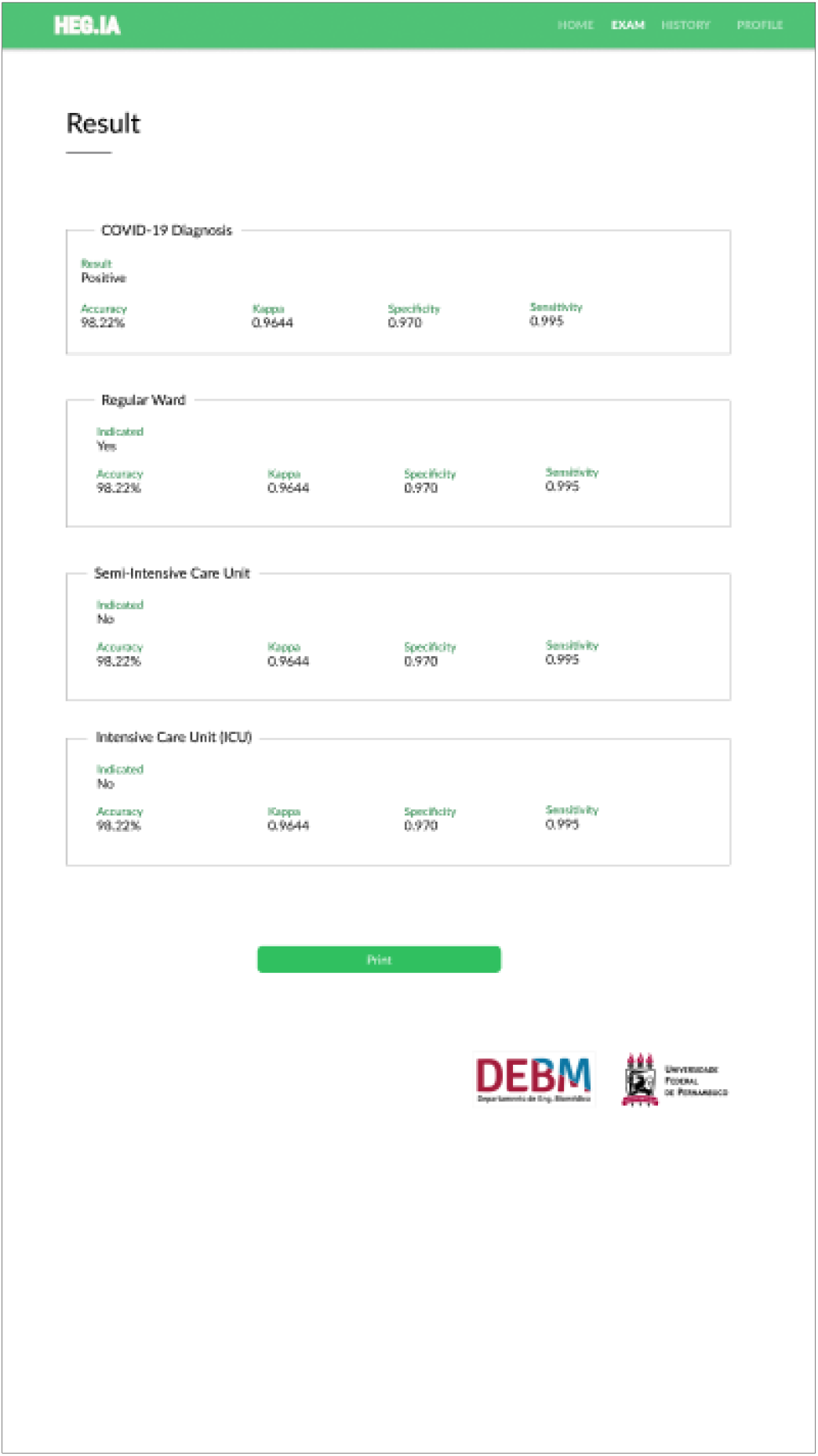
Results screen: In this screen it is possible to view the patient’s diagnostic report. In the report, the diagnosis for Covid-19 is available, as well as the hospitalization predictions, indicating whether the patient should be admitted to the regular ward, semi-intensive care unit, or to the ICU. Information on accuracy, kappa index, sensitivity and specificity of the determination of each of these scenarios are also available, in order to assist the physician’s decision making. In addition to viewing the report, it is also possible to print it.

## 5. Discussion

For the experiments comparing different classification models, Figures 4, 5, 6 and 7 show that, overall, Random Forest method overcame the others in both accuracy and kappa statistic. When using 90 trees in the Random Forest, the method achieved great results for all scenarios, always reaching accuracy above 90% and kappa statistic above 0.80. From these results we also found that the task of diagnosing SARS-Cov2 from the used blood tests (Figure 3) showed to be harder than indicating the type of hospitalization using the same group of exams.

For SARS-Cov2 detection scenario, all tested configurations of Random Forest and MLP showed good performance, with accuracies above 90% and kappa above 0.80. Less satisfying results were achieved by the SVMs, Bayesian networks and Random Tree. These last classifiers reached accuracy values between 80% and 90% with kappa statistic varying from 0.60 to 0.80. As for data dispersion, it was slightly greater for kappa results, when compared to accuracy. However, both graphs show low dispersion, indicating good reliability of the decision of all algorithms. The dispersion of Bayesian networks and Random Tree were slightly bigger than the other methods.

When regarding to classifiers performance on indicating the type of hospitalization, we found some outstanding results, specially using Random Forest with 90 and 100 trees. These classifiers achieved similar results for all three types of hospitalization. However, Random Forest with 90 trees performed slightly better for Regular Ward indication. In this scenario, once more, we achieve great results using MLP and Random Forest, with all results around 100% for accuracy and around 1.00 for kappa statistic. Impressive results were also reached using SVM with linear kernel, Naive Bayes network and Random Tree, all similar to the results using Random Forest and MLP. SVM with polynomial kernel and Bayes network performed worse than the other methods, showing higher data dispersion but still achieved accuracy values above 96% and kappa above 0.90. The data dispersion for the other method were very low, reaching its lowest values for Random Forest with 90 and 100 trees.

From the previously mentioned results, we found that the best classifier to solve both SARS-Cov2 detection and hospitalization indication problems was the Random Forest with 90 trees. This decision was supported by the excellent results shown on Tables 4 and 5. These table shows the method performance for the most relevant metrics regarding to diagnosis quality. For all scenarios described in both tables, we found accuracy, kappa, recall, sensitivity, precision, specificity and area under ROC curve really close to the their maximum values. These results indicate that the system showed a great overall efficiency. Considering this performance, we chose to use this model to build our HegIA Web Application.

## 6. Conclusion

The disease caused by the new type of coronavirus, the Covid-19, has posed a global public health challenge. Since this virus has a stronger human-to-human transmission ability, it has already led to millions of infected people and thousands of deaths since the beginning of the outbreak in December 2019. As pointed out by the World Health Organization, testing is our currently best strategy to fight Covid-19 pandemic spread.

The ground-truth test in Covid-19 diagnosis is the Reverse Transcription Polymerase Chain Reaction (RT-PCR) with DNA sequencing and identification. RT-PCR is precise, but takes several hours to be assessed. Another type of test, based on IgM/IgG antibodies, delivers results quickly, however they are nonspecific for Covid-19, and may have very low sensitivity and specificity. IgM/IgG tests do not directly detect the SARS-Cov2 presence, indeed they detect the serological evidence of recent infection. Considering this, the development of a diagnosis support system to provide fast results with high sensitivity and specificity is necessary and urgent. In this context, blood tests have some advantages. First, they are commonly used during medical screening. Besides that blood tests are less expensive and less time-consuming than other diagnosis methods. Thus providing a more accessible system.

In this study, we developed a web system, Heg.IA, which seeks to optimize the diagnosis of Covid-19 by combining blood tests, arterial gasometry results and artificial intelligence. From the system, a healthcare professional may have a diagnostic report after providing 41 hematological parameters from common blood tests. The system will indicate if the patient is infected with SARS-Cov2 virus, and also recommend the type of hospitalization (regular ward, semi-ICU, or ICU).

The proposed system is based on decision trees and achieved great performance of accuracy, kappa statistic, sensitivity, precision, specificity and area under ROC for all tested scenarios. Considering SARS-Cov2 detection, the system may play an important role as a highly efficient rapid test. The hospitalization recommendation module of the system can be important to speed up and improve decision-making regarding the referral of each patient.

Heg.IA application may be a way to overcome the testing unavailability in the context of this pandemic. Moreover, the hospitalization prediction can also support decision regarding to patient referral according to the hospitalization conditions of each health institution. Since the system is flexible and available online, we expect to reach different nations of the globe, specially less-favored countries and communities, in which the absence of testing is even more critical. Finally, we hope the system will provide wide access to Covid-19 effective diagnosis and thereby reach and help saving as many people as possible.

## Data Availability

Data is available under requirement.

## Acknowledgements

The authors are grateful to the Federal University of Pernambuco, Google Cloud COVID-19 Research Grant, and the Brazilian research agencies CAPES and CNPq, for the partial financial support of this research.

## Compliance with Ethical Standards

This study was funded by the Federal University of Pernambuco, Google Cloud COVID-19 Research Grant, and the Brazilian research agencies CAPES and CNPq.

All procedures performed in studies involving human participants were in accordance with the ethical standards of the institutional and/or national research committee and with the 1964 Helsinki declaration and its later amendments or comparable ethical standards.

